# Cardiac Structural and Functional Characteristics in Patients with Coronavirus Disease 2019: A Serial Echocardiographic Study

**DOI:** 10.1101/2020.05.12.20095885

**Authors:** Ge Heng, Zhu Mingli, Du Jing, Zhou Yong, Wang Wei, Zhang Wei, Jiang Handong, Qiao Zhiqing, Gu Zhichun, Li Fenghua, Pu Jun

**Affiliations:** State Key Laboratory of Oncogenes and Related Genes, Department of Cardiology, Department of Ultrasound, Department of Respiratory, Department of Pharmacy, Renji Hospital, School of Medicine, Shanghai Jiaotong University, Shanghai 200127, China

## Abstract

**BACKGROUND:** Increasing attention has been paid to cardiac involvement in patients with coronavirus disease 2019 (COVID-19). Yet, scarce information is available regarding the morphological and functional features of cardiac impairments in these patients.

**METHODS:** We conducted a prospective and serial echocardiographic study to investigate the structural and functional cardiac changes among COVID-19 patients admitted to the intensive care unit (ICU). From January 21 to April 8, 2020, a total of 51 ICU patients (31 critically ill and 20 severely ill) with confirmed COVID-19 were monitored by serial transthoracic echocardiography examinations. Outcomes were followed up until April 8, 2020.

**RESULTS:** Of 51 ICU patients, 33 (64.7%) had cardiovascular comorbidities. Elevations of levels of cardiac biomarkers including high-sensitivity cardiac troponin-I (hs-cTnI) and brain natriuretic peptide were observed in 62.7% and 86.3% of patients, respectively. Forty-two (82.3%) had at least one left-heart and/or right-heart echocardiographic abnormality. The overall median left ventricular ejection fraction (LVEF) was 65.0% (IQR 58.0–69.0%), with most (44/86.3%) having preserved LVEF. Sixteen patients (31.4%) had increased pulmonary artery systolic pressure, and 14 (27.5%) had right-ventricle (RV) enlargement. During the study period, 12 (23.5%) patients died. LVEF was comparable between survivors and non-survivors, while non-survivors had more often pulmonary hypertension (58.3% vs. 23.1%; *P*=0.028) and RV enlargement (58.3% vs. 17.9%, *P*=0.011). Kaplan-Meier analysis demonstrated similar survival curves between patients with vs. without echocardiographic left-heart abnormalities (*P*=0.450 by log-rank test), while right-heart abnormalities had adverse impact on mortality (*P*=0.012 by log-rank test).

**CONCLUSIONS:** Typical cardiac abnormality in ICU patients with COVID-19 was right-heart dysfunction with preserved LVEF. Echocardiographic right-heart dysfunction was associated with disease severity and increased mortality in patients affected by COVID-19.

**CLINICAL TRIAL REGISTRATION:** Unique identifier: NCT04352842.

## INTRODUCTION

The ongoing pandemic of coronavirus disease 2019 (COVID-19) caused by severe acute respiratory syndrome coronavirus 2 (SAR-CoV-2) has had many casualties ^1^. Although an acute respiratory syndrome is the main manifestation of the disease ^2^, understanding the impact of the virus on other organs is of importance. Multiple organ dysfunction/failure, including cardiovascular manifestations, has contributed to increased mortality in patients with COVID-19. Some studies have reported cardiac injury as evidenced by elevated levels of biomarkers such as high-sensitivity cardiac troponin-I (hs-cTnI) and brain natriuretic peptide (BNP) ^2-4^ Cardiac complications in several case reports have included acute heart failure, cardiac rupture, and sudden cardiac arrest ^5-7^ However, autopsy studies have not found evidence of direct myocyte assault by the virus or apparent cardiac myocyte necrosis/apoptosis ^8^. Instead, typical pathological findings have included mononuclear inflammatory infiltration in the myocardial interstitium without substantial cardiomyocytes damage ^9,10^. Thus, a significant gap exists in our knowledge between clinical observations and postmortem findings. At present, the in vivo morphological and functional features of cardiac injury remain unknown.

The present prospective and serial echocardiographic study aimed to investigate the cardiac structural and functional changes in patients with COVID-19 who were admitted to an intensive care unit (ICU), and to compare cardiac characteristics between deceased and surviving patients.

## METHODS

### Study Design and Participants

From January 21, 2020 to April 8, 2020, we conducted a prospective observational study to investigate the cardiac structural and functional characteristics by transthoracic echocardiographic examinations in a cohort of patients who were admitted to our ICU for COVID-19 (Trial Registration: NCT04352842). Including criteria were patients who: 1) had been diagnosed with COVID-19 according to the criteria established by WHO interim guidance ^2,11,12^ and 2) were admitted to the ICU because of severe or critical conditions. Patients who were < 18 years old or those whose entire stay in hospital lasted for < 48 hours were excluded. The severity of the disease was categorized according to published criteria ^13,14^ Criteria for severe cases included any of the following: 1) Respiratory rate ≥30 per minute; 2) blood oxygen saturation (SPO2) ≤ 93% at rest; 3) partial pressure of arterial oxygen to fraction of inspired oxygen ratio <300; or 4) more than 50% of lung infiltrates within 24 to 48 hours. Patients needing mechanical ventilatory support or presenting with septic shock or multiple organ dysfunction or failure constituted the critical cases.

The study protocol conformed to the Ethical Guidelines of the 1975 Declaration of Helsinki. All identification information that would impact personal privacy was removed during data collection. The Hospital Ethics Commission approved the study protocol and waived written informed consent for emerging infectious diseases according to the regulation issued by the National Health Commission of the People’s Republic of China.

### Data Collection

All epidemiological, clinical, laboratory, and outcome parameters were prospectively collected with standardized data collection forms from an electronic medical records system. Personal history was confirmed with patients or family members. Two researchers (H.G. and M.Z.) independently reviewed the forms to double check the data collected.

### Echocardiography Protocol and Data Analysis

Echocardiography was performed using a GE Vingmed Ultrasound System (GE Healthcare, Horten, Norway) with special protection in the contaminated area. Two-dimensional (2D) and Doppler echocardiographic measurements followed the recommendations of the American Society of Echocardiography ^15^. Protections of the echocardiographer and cleaning of the equipment were in accordance with the latest ESC recommendations ^16^. Conventional parasternal long-axis view as well as apical 4-chamber, 2-chamber and 3-chamber views were obtained. Images were analyzed using GE Echopac commercial software. All echocardiography determinations were based on the observations of two independent experienced reviewers (J.D. and W.W.) who were blinded to other data.

#### Determination of Left-heart Parameters

Left atrial (LA) anteroposterior diameter, left ventricular (LV) end-diastolic diameter (LVEDD), LV end-systolic diameter (LVESD), and interventricular septum (IVS) thickness were measured in the parasternal long-axis view using M-mode echocardiography and 2D echocardiography. LA anteroposterior dimension >40mm in male or >38mm in female patients defined LA enlargement. LVEDD >58mm in male or >52mm in female patients defined LV enlargement. IVS thickness >10mm defined IVS hypertrophy. LV ejection fraction (LVEF) was calculated with the use of the modified Simpson’s rule in apical 4-chamber and 2-chamber views. Mitral E/A ratio abnormality was an E/A ratio <1 (mitral inflow pattern) in the apical 4-chamber view. Color-coded TDI by apical 4-chamber view was used to determine the mean early and late velocities at both septal and lateral mitral annuli, and average E/e’ ratio > 14 was considered as abnormal. Mitral regurgitation was categorized as mild, moderate or severe according to the width of the vena contracta.

#### Determination of Right-heart Parameters

Right atrial (RA) diameter was measured in the apical four-chamber view as the distance between the lateral RA wall and interatrial septum. RA diameter >44mm defined RA enlargement. Right ventricular (RV) basal diameter was measured in the RV-focused view. RV basal diameter >41mm defined RV enlargement. Pulmonary artery systolic pressure (PASP) was estimated by tricuspid regurgitation pressure gradient, and PASP >40mmHg defined increased pulmonary artery pressure.

Tricuspid annular plane systolic excursion (TAPSE) was measured by M-mode echocardiography with the cursor aligned along the direction of the tricuspid lateral annulus in the apical 4-chamber view. TAPSE <17 mm defined RV systolic dysfunction. Tricuspid regurgitation was categorized as mild, moderate, or severe according to the width of the vena contracta.

### Events and Follow-up

Clinical outcomes were censored at the time of data cutoff which occurred on April 8, 2020. For patients who were discharged before April 8, follow-up was continued by telephone interview. Time from diagnosis to death was recorded.

### Statistics

Data analyses were performed using SPSS 23.0 (SPSS Inc., Chicago, Illinois) and SAS version 9.2 (SAS Institute, Cary, North Carolina). Categorical data are expressed as absolute values and percentages and were compared using chi-square or Fisher exact tests, and continuous data are reported as median (interquartile range [IQR]) and were compared using Kruskal-Wallis/Wilcoxon rank sum tests. Cox proportional hazards regression model was used to estimate hazard ratios (HRs) and 95% confidence intervals (CIs) with adjustment for potential confounders for mortality risk. Multivariable Cox regression models were used to determine the independent risk factors for death. Two criteria were considered necessary for a variable to be entered in the multivariable analysis model: (1) a univariate *P* value for survivors vs. non-survivors comparison ≤0.10; and (2) a plausible association with the risk of death in COVID-19 according to data provided by the literature. Time to events was described by the Kaplan-Meier curves, and groups were compared using the log-rank test. Inferential statistical tests were conducted at a significance level of 0.05. The authors had full access to and take full responsibility for the integrity of the data. All authors have read and agreed to the report as written.

## Results

### Demographics and Baseline Characteristics

From Jan 21, 2020, to April 8, 2020, a total of 51 patients who were admitted to the ICU and underwent serial echocardiography examinations were prospectively enrolled in this study. All patients had laboratory-confirmed COVID-19 infection. Summary of general information of all study patients is shown in Supplementary Table 1.

The median patient age was 70 years (IQR, 58.0–79.0; range, 25-93 years); and 37 (72.5%) were male. Thirty-three (64.7%) had underlying cardiovascular diseases including hypertension (43.1%), coronary artery disease (23.5%), congestive heart failure (7.8%), and atrial fibrillation (5.9%); and 36 (70.6%) had other comorbidities including diabetes mellitus (31.4%), pulmonary disease (13.7%), renal disease (23.5%), liver disease (11.8%), or cancer (9.8%). Overall, 84.3% had lymphopenia, 33.3% had monopenia, 92.2% had increased D-Dimer levels, and 45.1% had increased fibrin degradation product (FDP) levels. Hyperinflammatory status was observed in most patients as evidenced by increased peak concentrations of high-sensitivity C-reactive protein (hs-CRP) (84.3%) and interleukin-6 (IL-6) (80.4%). Elevated levels of cardiac markers including hs-cTnI and BNP were observed in 62.7% and 86.3% of patients, respectively.

According to the severity of the disease ^13,14^, 31 (60.8%) were classified as critically ill and 20 patients (39.2%) were classified as severely ill. Demographic variables including the prevalence of underlying cardiovascular diseases and other comorbidities were similarly distributed between severely ill with critically ill patients (Table 1). However, compared with severely ill patients, critically ill patients had higher blood leukocyte count (*P*=0.002) and neutrophilic granulocyte percentage (*P*<0.001), as well as lower lymphocyte percentage (*P*=0.001), monocyte percentage (*P*=0.001), and platelet count (*P*=0.002). Compared with severely ill patients, the levels of D-Dimer (*P*=0.008) and FDP (*P*=0.035), inflammation markers including hs-CRP (*P*<0.001) and IL-6 (*P*<0.001), and cardiac injury indicators including hs-cTnI (*P*<0.001) and BNP (*P*=0.001), were greatly increased in critically ill patients.

**Table 1.** Demographics and baseline characteristics classified by disease severity (critically ill patients vs. severely ill patients)

| Characteristics | All patients (n=51) | Critically ill patients (n=31) | Severely ill patients (n=20) | *P* value |
| --- | --- | --- | --- | --- |
| Age, years | 70 (58-79) | 72 (62-80) | 66 (55.5-76.75) | 0.434 |
| Sex |  |  |  | 0.257 |
| Men, n/% | 37 (72.5) | 24 (77.4) | 13 (65.0) |  |
| Women, n/% | 14 (27.5) | 7 (22.6) | 7 (35.0) |  |
| Current smoking, n/% | 7 (13.7) | 4 (12.9) | 3 (15.0) | 0.571 |
| Any comorbidity, n/% | 42 (82.4) | 28 (90.3) | 14 (70) | 0.070 |
| Cardiovascular diseases, n/% | 33 (64.7) | 22 (71.0) | 11 (55.0) | 0.193 |
| -Hypertension, n/% | 22 (43.1) | 12 (38.7) | 10 (50.0) | 0.306 |
| -Coronary artery disease, n/% | 12 (23.5) | 10 (32.3) | 2 (10.0) | 0.065 |
| -Previous myocardium infarction, n/% | 4 (7.8) | 4 (12.9) | 0 (0) | 0.126 |
| -Atrial fibrillation, n/% | 3 (5.9) | 3 (9.7) | 0 (0) | 0.216 |
| -Heart failure, n/% | 4 (7.8) | 3 (9.7) | 1 (5.0) | 0.485 |
| -Stroke, n/% | 7 (13.7) | 4 (12.9) | 3 (15.0) | 0.571 |
| Diabetes mellitus, n/% | 16 (31.4) | 10 (32.3) | 6 (30.0) | 0.559 |
| Pulmonary disease, n/% | 7 (13.7) | 5 (16.1) | 2 (10.0) | 0.429 |
| Renal disease, n/% | 12 (23.5) | 6 (19.4) | 6 (30.0) | 0.293 |
| Liver disease, n/% | 6 (11.8) | 3 (9.7) | 3 (15.0) | 0.438 |
| Malignancy, n/% | 5 (9.8) | 3 (9.7) | 2 (10.0) | 0.660 |
| Laboratory findings |  |  |  |  |
| -Leukocytes*10<sup>9</sup>/L | 7.82 (5.67-12.60) | 9.82 (7.35-14.00) | 6.11 (4.88-8.40) | 0.002 |
| -Erythrocytes*10<sup>12</sup>/L | 3.28 (2.88-3.97) | 3.24 (2.63-3.24) | 3.66 (2.99-4.16) | 0.153 |
| -Hemoglobin, g/L | 105.00 (80.00-119.00) | 105.00 (79.00-115.00) | 106.50 (94.25-128.75) | 0.153 |
| -Neutrophil, % | 83.30 (73.50-91.40) | 90.40 (82.00-92.70) | 74.40 (68.43-82.38) | <0.001 |
| -Lymphocyte, % | 7.80 (0.51-14.60) | 1.30 (0.34-8.90) | 13.50 (7.83-23.60) | 0.001 |
| -Monocytes, % | 5.80 (2.30-8.80) | 4.10 (1.50-6.40) | 8.80 (5.88-11.48) | 0.001 |
| -Platelets*10<sup>9</sup>/L | 170.00 (117.0-258.00) | 142.00 (91.00-204.00) | 209.00 (167.25-357.70) | 0.002 |
| -AST, u/L | 38.00 (19.00-65.00) | 44.00 (24.00-67.00) | 19.00 (16.00-54.00) | 0.015 |
| -ALT, u/L | 27.00 (15.00-68.00) | 32.00 (18.00-70.00) | 23.50 (14.00-39.50) | 0.171 |
| -Albumin, g/L | 31.30 (27.68-34.88) | 29.00 (26.82-32.67) | 32.45 (29.13-38.11) | 0.099 |
| -Creatinine, umol/L | 77.49 (53.30-138.10) | 79.51 (48.17-139.01) | 73.99 (58.88-136.95) | 0.787 |
| -BUN, mmol/L | 8.40 (5.00-13.70) | 9.40 (5.40-16.00) | 6.65 (4.79-10.18) | 0.105 |
| -D-Dimer, mg/L | 2.44 (1.16-5.04) | 4.10 (1.43-8.04) | 1.73 (0.77-2.73) | 0.008 |
| -FDP, mg/L | 4.40 (2.95-7.01) | 5.37 (3.69-13.53) | 3.95 (2.68-6.04) | 0.035 |
| -BNP, pg/mL | 674.04 (251.30-1406.00) | 1043.00 (355.74-1679.00) | 265.88 (69.27-748.07) | 0.001 |
| -Hs-cTnI, ng/ml | 0.07 (0.02-0.23) | 0.17 (0.06-0.32) | 0.028 (0.01-0.09) | <0.001 |
| -Hs-CRP, mg/L | 66.26 (20.99-105.79) | 95.66 (56.00-133.00) | 21.30 (6.81-59.08) | <0.001 |
| -Procalcitonin, ng/mL | 0.43 (0.10-2.04) | 0.92 (0.29-3.74) | 0.16 (0.07-0.43) | 0.003 |
| -IL-6 peak, pg/mL | 39.81 (10.00-674.38) | 250.00 (26.11-1582.69) | 12.09 (4.57-30.37) | <0.001 |
| -CD 4<sup>+</sup> T cell , /uL | 115.75 (63.00-311.00) | 83.74 (56.25-144.99) | 387.00 (203.54-540.00) | <0.001 |
| Oxygen treatments |  |  |  | <0.001 |
| -Nasal cannula, n/% | 13 (25.5) | 2 (6.5) | 11 (55.0) |  |
| -HFNC, n/% | 12 (23.5) | 3 (9.7) | 9 (45.0) |  |
| -NIV, n/% | 5 (9.8) | 5 (16.1) | 0 (0) |  |
| -IMV, n/% | 21 (41.2) | 21 (67.7) | 0 (0) |  |
| ECMO, n/% | 9 (17.6) | 9 (29.0) | 0 (0) | 0.007 |

Categorical data are expressed as absolute values and percentage and were compared using chi-square or Fisher exact tests, and continuous data are reported as median (interquartile range [IQR]) and were compared using Kruskal-Wallis/Wilcoxon rank sum tests. AST= Aspartate aminotransferase,
ALT=Alanine aminotransferase, BUN= Blood urea nitrogen, FDP= Fibrin degradation product, BNP= Brain natriuretic peptide, Hs-cTnI= hypersensitive troponin I, Hs-CRP= hypersensitive C-reactive protein, IL-6= Interleukin-6, HFNC= High-flow nasal cannula, NIV= Non-invasive mechanical ventilation, IMV= Invasive mechanical ventilation, ECMO=extracorporeal membrane oxygenation.

### Echocardiographic Findings

Forty-two (82.3%) had at least one abnormal left and/or right-heart echocardiographic parameter: 38 patients (74.5%) had at least one abnormal left heart echocardiographic manifestation, and 22 patients (43.1%) had at least one abnormal right heart echocardiographic manifestation. Detailed comparisons of echocardiographic manifestations between severely and critically ill patients have been illustrated in Figure 1 and Table 2.

**Figure 1.**
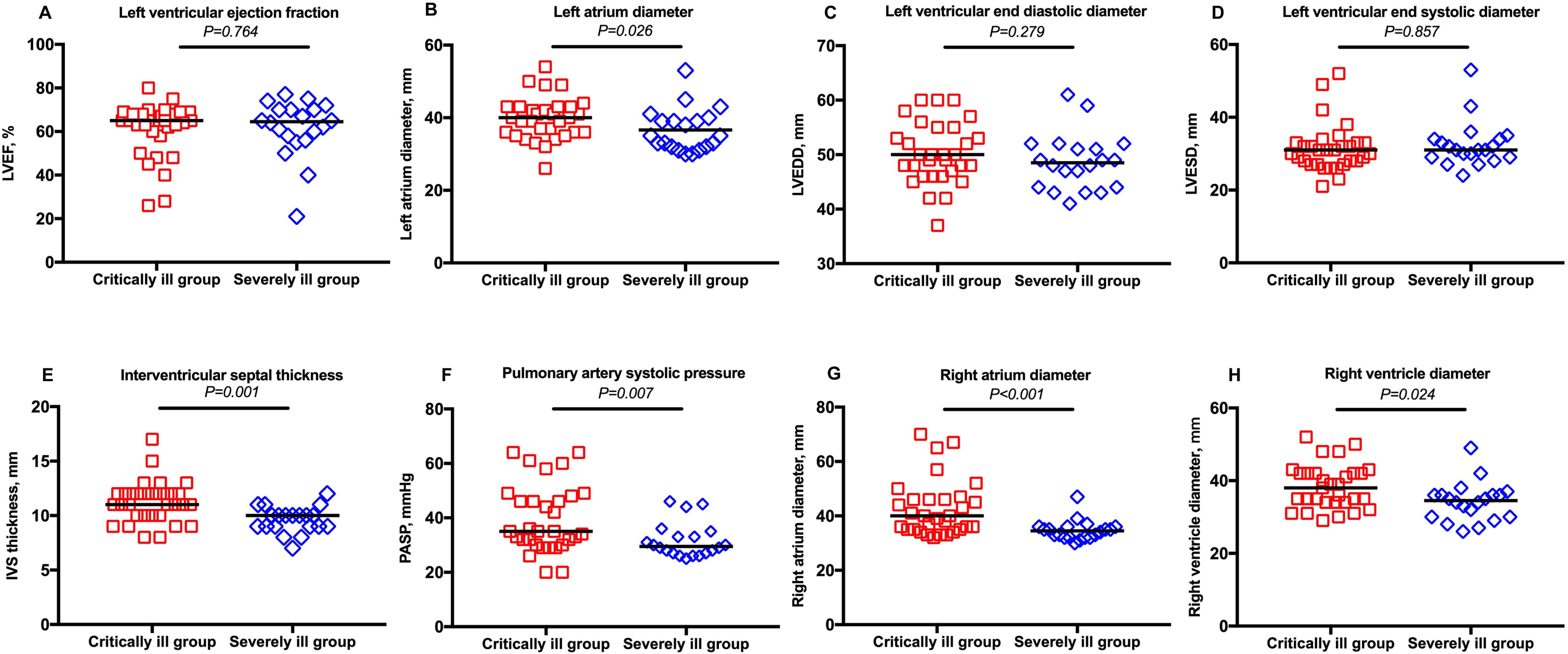
Comparison of cardiac structural and functional parameters between critically ill and severely ill COVID-19 patients. Categorical data were compared using chi-square or Fisher exact tests, and continuous data were compared using Kruskal-Wallis/Wilcoxon rank sum tests. LVEF, left ventricular eject fraction; LVESD, left ventricular end-systolic diameter; LVEDD, left ventricular end-diastolic diameter; IVS, interventricular septal thickness; PASP, pulmonary artery systolic pressure.

**Figure 2.**
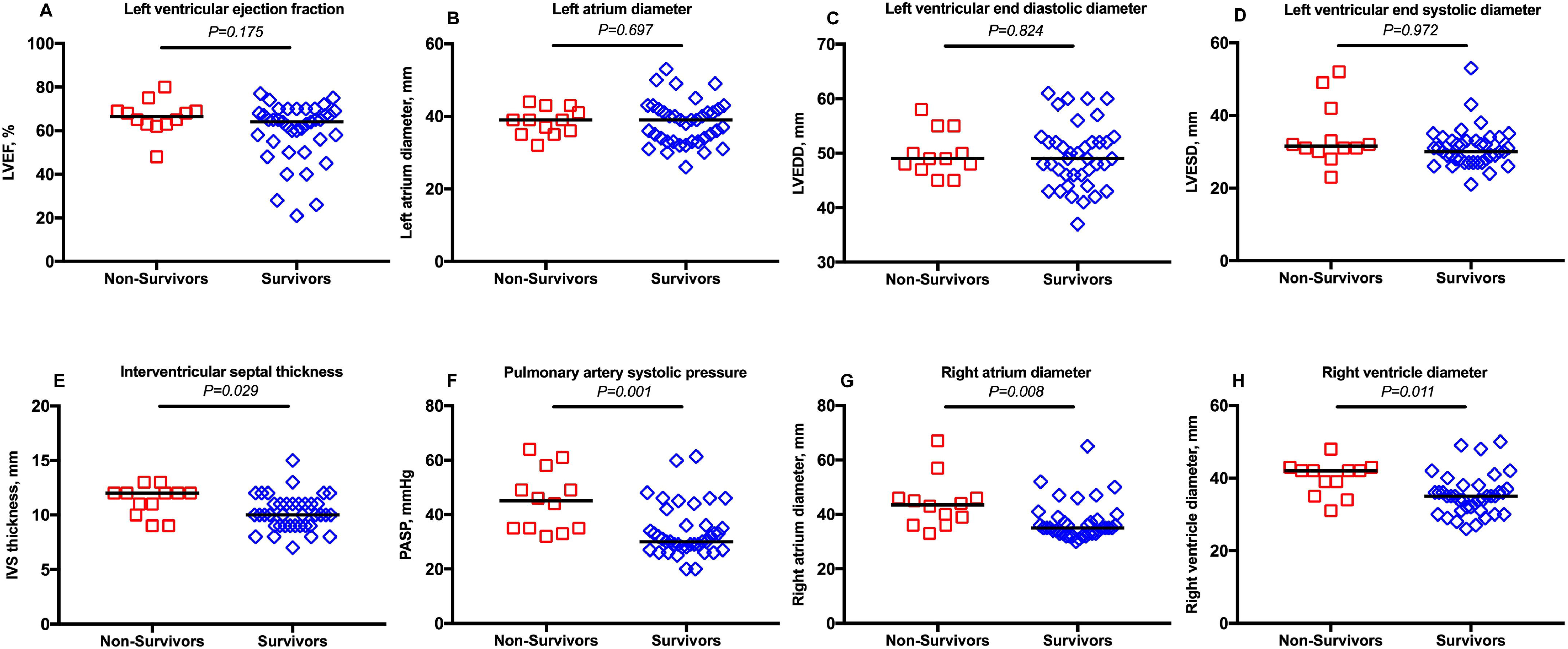
Comparison of cardiac structural and functional parameters between non-survivors and survivors. Categorical data were compared using chi-square or Fisher exact tests, and continuous data were compared using Kruskal-Wallis/Wilcoxon rank sum tests. LVEF, left ventricular eject fraction; LVESD, left ventricular end-systolic diameter; LVEDD, left ventricular end-diastolic diameter; IVS, interventricular septal thickness; PASP, pulmonary artery systolic pressure.

**Table 2.**
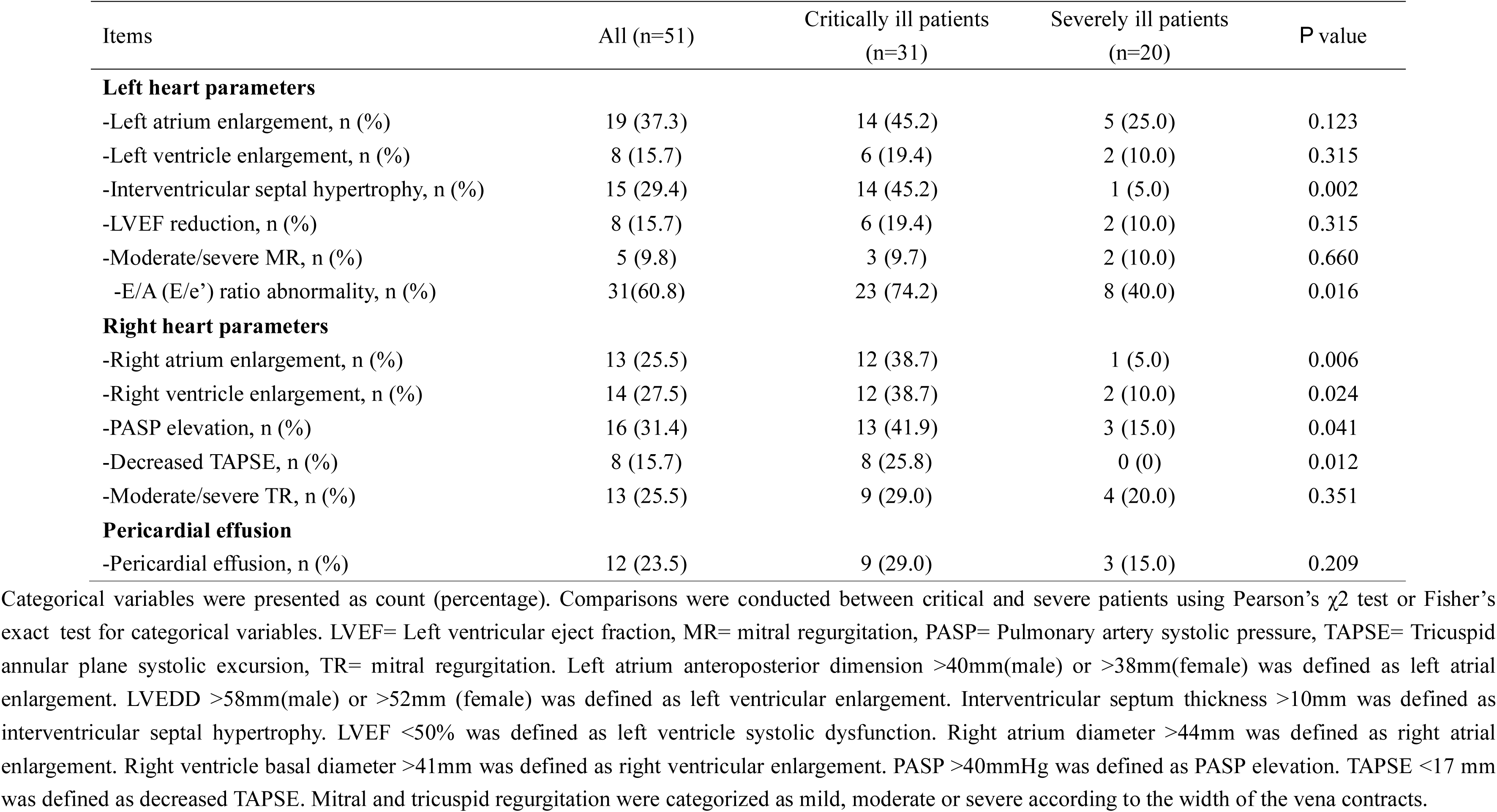
Echocardiographic characteristics classified by disease severity (critically ill patients vs. severely ill patients)

#### Structural and Functional Left Heart Changes

The overall median LVEF was 65.0% (IQR 58·0–69·0%), with no difference between critically ill (66.5%, IQR 63·0–69·0%) and severely ill (64.0%, IQR 55·0–69·0%; *P*=0.64) cases. Most patients (44/86.3%) presented preserved LVEF; and only 8 patients (15.7%) had reduced LVEF (<50%).

Almost all patients with reduced LVEF had history of cardiovascular diseases (previous myocardial infarction (MI) in 3, congestive heart failure in 4, and hypertension in 6). Regional wall motion abnormalities were observed only in patients with previous MI; and in two patients with previous MI and reduced LVEF, there was thrombus formation within an LV aneurysm. Eight patients (15.7%) had enlarged LV, and five (9.8%) presented with moderate/severe mitral regurgitation. LVEDD (*P*=0.279) and LVESD (*P*=0.857) were comparable between critically and severely ill patients.

Overall, 31 (60.8%) had E/A ratio and E/e’ ratio abnormality: 23 (74.2%) critically ill and 8 (40.0%) severelly ill patients (*P*=0.016). Nineteen (37.3%) presented LA enlargement, and critically ill patients had larger LA diameter (40.0 [IQR 36.0-43.0] mm vs. 35.0 [IQR 32.0-39.8] mm; *P*=0.026) and thicker IVS (11.0 [IQR 10.0-12.0] mm vs.10.0 [IQR 9.0-10.0] mm; *P*=0.001) versus severely ill patients.

#### Structural and Functional Right Heart Characteristics

Sixteen patients (31.4%) had increased PASP (>40mmHg): 13 (41.9%) were critically ill and three (15.0%) were severely ill (*P*=0.041). The prevalence of RV systolic dysfunction (TAPSE <17mm) was higher in critically ill vs. severely ill patients (8/25.8% vs. 0/0%, *P*=0.012) along with larger RA (*P*<0.001) and RV (*P*=0.024) diameters. The prevalence of moderate to severe tricuspid regurgitation was comparable (*P*=0.351).

#### Pericardial Effusion

Pericardial effusion was noted in 12 (23.5%) patients with median thickness of 8.0mm (IQR, 5.0–9.0mm; range, 1-11 mm), and similar prevalence between critically ill (9/29.0%) and severely ill (3/15.0%) cases (*P*=0.209).

### Cardiac Involvement in Survivors vs. Non-survivors

Twelve (23.5%) patients died. The median age of non-survivors was 75.0 years (IQR, 57.75-80.50 years; range, 25-82 years), and 75.0% were male. As shown in Table 3, the demographic characteristics and the prevalence of chronic diseases were comparable between survivors and non-survivors, with only a tendency of a more frequent cancer history in non-survivors (*P*=0.078). However, compared with survivors, non-survivors had higher neutrophilic granulocyte percentage (*P*=0.002), and aspartate aminotransferase (AST) (*P*=0.022), D-Dimer (*P*=0.024), FDP (*P*=0.044), hs-CRP (*P*=0.024), and IL-6 (P<0.001) levels as well as lower lymphocyte percentage (*P*=0.014), monocyte percentage (*P*=0.003), platelet count (P<0.001), and albumin level (*P*=0.015). Cardiac injury indicators such as hs-cTnI (*P*=0.039) and BNP (*P*=0.045) tended to increase in non-survivors (Table 3).

**Table 3.** Demographics and baseline characteristics classified by clinical outcome (non-survivors vs. survivors)

| Characteristics | All patients (n=51) | Non-survivors (n=12) | Survivors (n=39) | *P* value |
| --- | --- | --- | --- | --- |
| Age, years | 70 (58-79) | 75.00 (57.75-80.50) | 67.00 (58.00-78.00) | 0.649 |
| Sex |  |  |  | 0.572 |
| Men, n/% | 37 (72.5) | 9 (75.0) | 28 (71.8) |  |
| Women, n/% | 14 (27.5) | 3 (25.0) | 11 (28.2) |  |
| Current smoking, n/% | 7 (13.7) | 1 (8.3) | 6 (15.4) | 0.471 |
| Any comorbidity, n/% | 42 (82.4) | 10 (83.3) | 32 (82.1) | 0.646 |
| Cardiovascular diseases, n/% | 33 (64.7) | 6 (50.0) | 27 (69.2) | 0.190 |
| -Hypertension, n/% | 22 (43.1) | 6 (50.0) | 16 (41.0) | 0.412 |
| -Coronary artery disease, n/% | 12 (23.5) | 2 (16.7) | 10 (25.6) | 0.416 |
| -Previous myocardium infarction, n/% | 4 (7.8) | 0 (0) | 4 (10.3) | 0.329 |
| -Atrial fibrillation, n/% | 3 (5.9) | 2 (16.7) | 1 (2.6) | 0.134 |
| -Heart failure, n/% | 4 (7.8) | 0 (0) | 4 (10.3) | 0.329 |
| -Stroke, n/% | 7 (13.7) | 3 (25.0) | 4 (10.3) | 0.201 |
| Diabetes mellitus, n/% | 16 (31.4) | 2 (16.7) | 14 (35.9) | 0.186 |
| Pulmonary disease, n/% | 7 (13.7) | 2 (16.7) | 5 (12.8) | 0.529 |
| Renal disease, n/% | 12 (23.5) | 1 (8.3) | 11 (28.2) | 0.151 |
| Liver disease, n/% | 6 (11.8) | 3 (25.0) | 3 (7.7) | 0.134 |
| Malignancy, n/% | 5 (9.8) | 3 (25.0) | 2 (5.1) | 0.078 |
| Laboratory findings |  |  |  |  |
| -Leukocytes*10<sup>9</sup>/L | 7.82 (5.67-12.60) | 9.61 (6.77-16.38) | 7.35 (5.66-11.63) | 0.206 |
| -Erythrocytes*10<sup>12</sup>/L | 3.28 (2.88-3.97) | 3.14 (2.56-3.48) | 3.34 (2.96-4.08) | 0.115 |
| -Hemoglobin, g/L | 105.00 (80.00-119.00) | 90.00 (79.25-112.50) | 106.00 (85.00-128.00) | 0.186 |
| -Neutrophil, % | 83.30 (73.50-91.40) | 92.00 (79.25-112.50) | 80.95 (71.78-90.40) | 0.002 |
| -Lymphocyte, % | 7.80 (0.51-14.60) | 0.91 (0.30-7.00) | 8.50 (0.76-17.50) | 0.014 |
| -Monocytes, % | 5.80 (2.30-8.80) | 2.00 (1.30-4.33) | 6.40 (3.40-9.00) | 0.003 |
| -Platelets*10<sup>9</sup>/L | 170.00 (117.0-258.00) | 90.00 (41.75-114.75) | 190.00 (146.00-294.00) | <0.001 |
| -AST, u/L | 38.00 (19.00-65.00) | 62.50 (39.50-74.50) | 24.00 (19.00-57.00) | 0.022 |
| -ALT, u/L | 27.00 (15.00-68.00) | 52.00 (26.00-70.00) | 24.00 (15.00-68.00) | 0.183 |
| -Albumin, g/L | 31.30 (27.68-34.88) | 26.35 (25.70-31.50) | 32.00 (29.00-36.30) | 0.015 |
| -Creatinine, umol/L | 77.49 (53.30-138.10) | 56.00 (43.85-114.39) | 79.51 (62.80-139.01) | 0.140 |
| -BUN, mmol/L | 8.40 (5.00-13.70) | 10.59 (6.70-16.00) | 8.40 (4.72-12.23) | 0.155 |
| -D-Dimer, mg/L | 2.44 (1.16-5.04) | 4.10 (2.20-10.95) | 2.01 (0.99-4.38) | 0.024 |
| -FDP, mg/L | 4.40 (2.95-7.01) | 6.96 (4.24-41.92) | 4.29 (2.75-6.38) | 0.044 |
| -BNP, pg/mL | 674.04 (251.30-1406.00) | 1071.39 (436.82-1820.50) | 391.90 (164.00-1406.00) | 0.045 |
| -Hs-cTnI, ng/ml | 0.07 (0.02-0.23) | 0.20 (0.03-0.54) | 0.07 (0.02-0.18) | 0.039 |
| -Hs-CRP, mg/L | 66.26 (20.99-105.79) | 95.82 (74.92-114.95) | 42.67 (18.36-95.66) | 0.024 |
| -Procalcitonin, ng/mL | 0.43 (0.10-2.04) | 1.53 (0.56-3.32) | 0.29 (0.10-1.83) | 0.087 |
| -IL-6 peak, pg/mL | 39.81 (10.00-674.38) | 1071.39 (463.82-1820.50) | 19.48 (7.43-68.48) | <0.001 |
| -CD 4<sup>+</sup> T cell , /uL | 115.75 (63.00-311.00) | 67.21 (50.55-118.68) | 193.00 (85.17-386.50) | 0.020 |
| Oxygen treatments |  |  |  | <0.001 |
| -Nasal cannula, n/% | 13 (25.5) | 0 (0) | 13 (33.3) |  |
| -HFNC, n/% | 12 (23.5) | 0 (0) | 12 (30.8) |  |
| -NIV, n/% | 5 (9.8) | 3 (25.0) | 2 (5.1) |  |
| -IMV, n/% | 21 (41.2) | 9 (75.0) | 12 (30.8) |  |
| ECMO, n/% | 9 (17.6) | 5 (41.7) | 4 (10.3) | 0.024 |

Categorical data are expressed as absolute values and percentage and were compared using chi-square or Fisher exact tests, and continuous data are reported as median (interquartile range [IQR]) and were
compared using Kruskal-Wallis/Wilcoxon rank sum tests. AST= Aspartate aminotransferase, ALT=Alanine aminotransferase, BUN= Blood urea nitrogen, FDP= Fibrin degradation product, BNP= Brain natriuretic peptide, Hs-cTnI= hypersensitive troponin I, Hs-CRP= hypersensitive C-reactive protein, IL-6= Interleukin-6, HFNC= High-flow nasal cannula, NIV= Non-invasive mechanical ventilation, IMV= Invasive mechanical ventilation, ECMO=extracorporeal membrane oxygenation.

When comparing echocardiographic findings, we did not observe any significant in left-heart parameters except for a thicker IVS in non-survivors (12.00 [IQR 10.00-12.00] mm vs.10.00 [IQR 9.00-11.00] mm; *P*=0.029). However, non-survivors had more abnormal right-heart parameters including elevated PASP (58.3% vs. 23.1%; *P*=0.028), RA enlargement (50.0% vs. 17.9%; *P*=0.036), RV enlargement (58.3% vs. 17.9%, *P*=0.011), and decreased TAPSE (41.7% vs. 7.7%, *P*=0.012) (Table 4).

**Table 4.** Echocardiographic characteristics in patient groups classified by clinical outcome (non-survivors vs. survivors)

| Items | All (n=51) | Non-survivors (n=12) | Survivors (n=39) | *P* value |
| --- | --- | --- | --- | --- |
| **Left heart parameters** |  |  |  |  |
| -Left atrium enlargement, n (%) | 19 (37.3) | 4 (33.3) | 15 (38.5) | 0.514 |
| -Left ventricle enlargement, n (%) | 8 (15.7) | 2 (16.7) | 6 (15.4) | 0.614 |
| -Interventricular septal hypertrophy, n (%) | 15 (29.4) | 7 (58.3) | 8 (20.5) | 0.018 |
| -LVEF reduction, n (%) | 8 (15.7) | 1 (8.3) | 7 (17.9) | 0.386 |
| -Moderate/severe MR, n (%) | 5 (9.8) | 2 (16.7) | 3 (7.7) | 0.335 |
| -E/A (E/e') ratio abnormality, n (%) | 31 ( 60.8 ) | 7 ( 58.3 ) | 24 ( 61.5 ) | 0.550 |
| **Right heart parameters** |  |  |  |  |
| -Right atrium enlargement, n (%) | 13 (25.5) | 6 (50.0) | 7 (17.9) | 0.036 |
| -Right ventricle enlargement, n (%) | 14 (27.5) | 7 (58.3) | 7 (17.9) | 0.011 |
| -PASP elevation, n (%) | 16 (31.4) | 7 (58.3) | 9 (23.1) | 0.028 |
| -Decreased TAPSE, n (%) | 8 (15.7) | 5 (41.7) | 3 (7.7) | 0.012 |
| -Moderate/severe TR, n (%) | 13 (25.5) | 4 (33.3) | 9 (23.1) | 0.358 |
| **Pericardial effusion** |  |  |  |  |
| -Pericardial effusion, n (%) | 12 (23.5) | 4 (33.3) | 8 (20.5) | 0.291 |

Categorical variables were presented as count (percentage). Comparisons were conducted between critical and severe patients using Pearson's $\chi^2$ test or Fisher's exact test for categorical variables. LVEF= Left ventricular eject fraction, MR= mitral regurgitation, PASP= Pulmonary artery systolic pressure, TAPSE= Tricuspid annular plane systolic excursion, TR= mitral regurgitation. Left atrium anteroposterior dimension >40mm(male) or >38mm(female) was defined as left atrial enlargement. LVEDD >58mm(male) or >52mm (female) was defined as left ventricular enlargement. Interventricular septum thickness >10mm was defined as interventricular septal hypertrophy. LVEF <50% was defined as left ventricle systolic dysfunction. Right atrium diameter >44mm was defined as right atrial enlargement. Right ventricle basal diameter >41mm was defined as right ventricular enlargement. PASP >40mmHg was defined as PASP elevation. TAPSE <17 mm was defined as decreased TAPSE. Mitral and tricuspid regurgitation were categorized as mild, moderate or severe according to the width of the vena contracts.

Figure 3 shows the serial changes in cardiac structural and functional parameters in 12 non-survivors. Compared with the echocardiographic findings on admission to the ICU, there was no difference in LVEF (63.50 [IQR 60.75-67.25]% vs. 64.50 [62.00-67.75]%; *P*=0.130), LVEDD (45.50 [43.00-49.75] mm vs. 48.50 [45.50-50.75] mm; *P*=0.777), and LVESD (31.00 [28.00-32.75] mm vs. 31.00 [27.50-33.75] mm, *P*=0.118) in non-survivors on the day before death. However, IVS thickness (9.50 [IQR 9.0-10.75] mm vs.12.0 [IQR 10.25-12.0] mm; *P*=0.001), PASP (31.00 [IQR 29.25-35.75] mmHg vs. 44.50 [33.50-50.50] mmHg; *P*=0.009), and RV diameter (35.00 [33.00-36.00] mm vs. 42.0 [33.25-42.75] mm; *P*=0.004) increased significantly in non-survivors before death.

**Figure 3.**
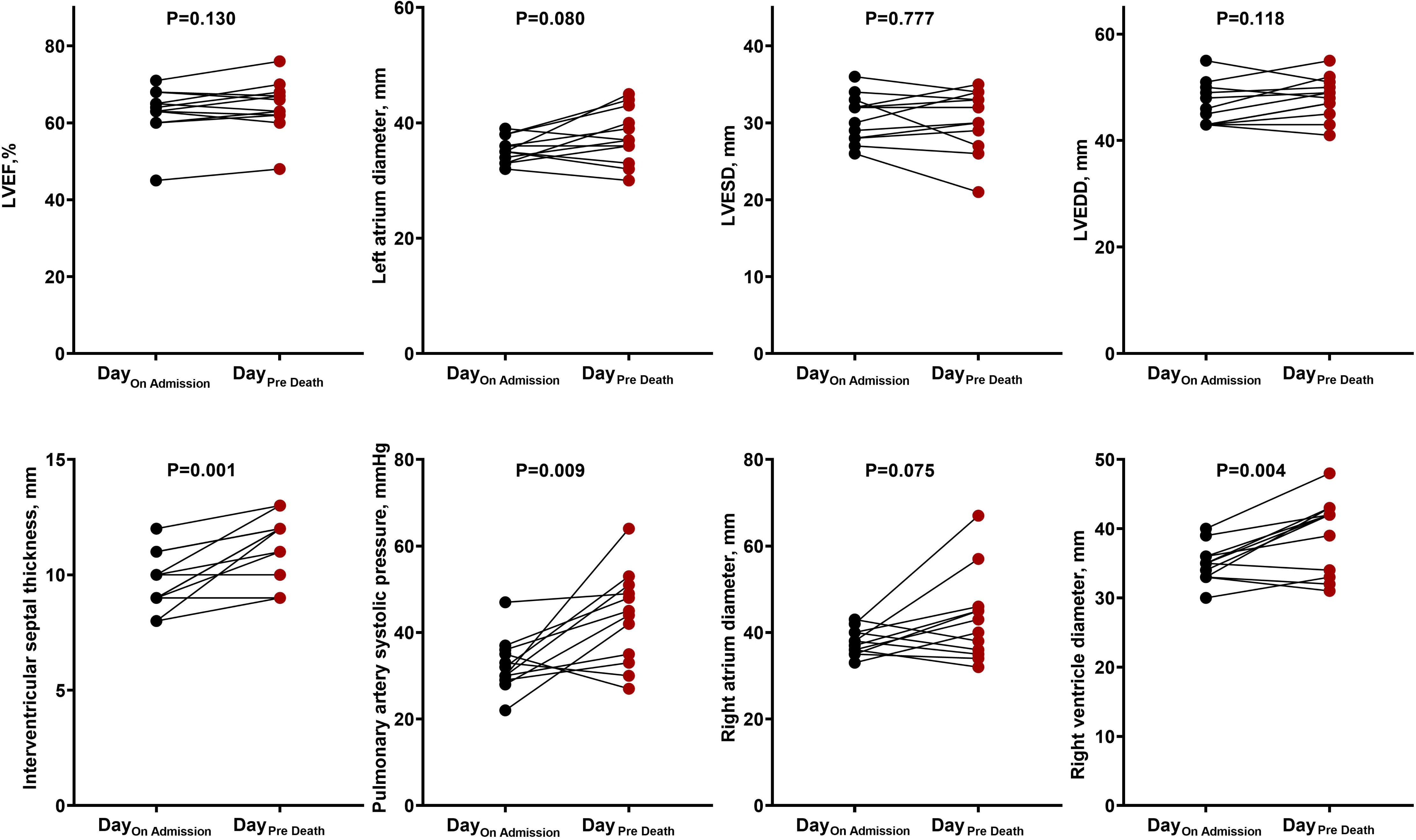
Changes in cardiac structural and functional parameters among 12 non-survivors. LVEF, left ventricular eject fraction; LVESD, left ventricular end-systolic diameter; LVEDD, left ventricular end-diastolic diameter.

Cox proportional hazards regression model was used to estimate the mortality risk for left-heart abnormality or right-heart abnormality with adjustment for potential confounders. Left-heart abnormality was not associated with mortality (HR 1.78 (95% CI 0.39 to 8.16), *P*_trend_=0.456. However, abnormalities of the right heart were associated with increased mortality in patients with COVID-19: HR (95% CI) for mortality was 4.58 (1.24 to 16.96), *P*_trend_^=^0.023, for patients with right-heart abnormality vs. patients without right-heart abnormality. As shown in Figure 4, Kaplan-Meier survival curves were similar between patients with vs. without left-heart abnormalities (*P*=0.450 by the log-rank test). However, when patients manifested right-heart abnormalities, the survival rate decreased (*P*=0.012 by the log-rank test).

**Figure 4.**
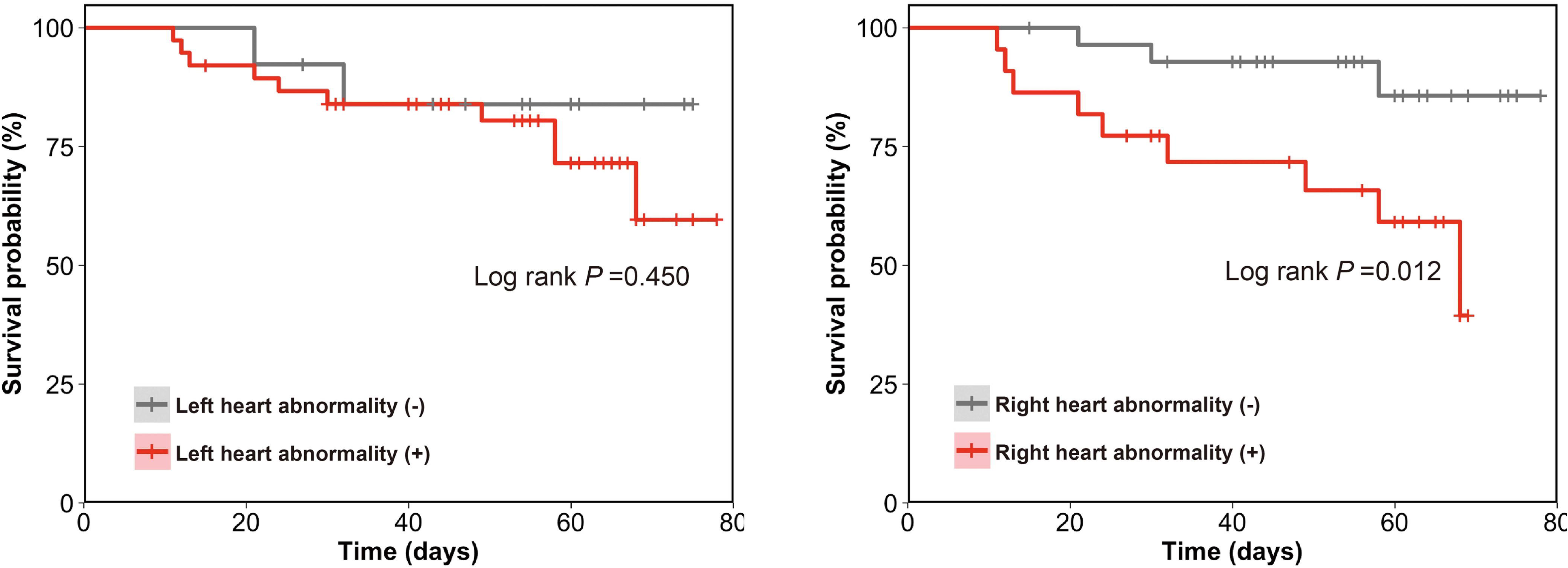
Impact of left-heart or right-heart echocardiographic abnormalities on mortality in COVID-19 patients during the study period. Time to events was described by the Kaplan-Meier curves, and groups were compared using the log-rank test.

### Impact of Use of Extracorporeal Membrane Oxygenation (ECMO)

Nine (17.6%) patients received ECMO. We monitored dynamic changes in cardiac structural and functional parameters before and after ECMO (Figure 5). ECMO reduced PASP (34.00 [IQR 30.50-40.50] mmHg vs. 25.00 [21.00-28.00] mmHg; *P*=0.011), and RA diameter (40.00 [IQR 37.00-50.00] mm vs. 36.00 [IQR 33.50-38.00] mm; *P*=0.044) and RV (40.50 [33.25-45.75] mm vs. 35.00 [34.00-39.50] mm; *P*=0.054) in critically ill cases. Of 9 patients receiving ECMO, four survived and five died. The changes in echocardiographic parameters post-ECMO in survivors vs. non-survivors are illustrated in Supplementary Figure 1.

**Figure 5.**
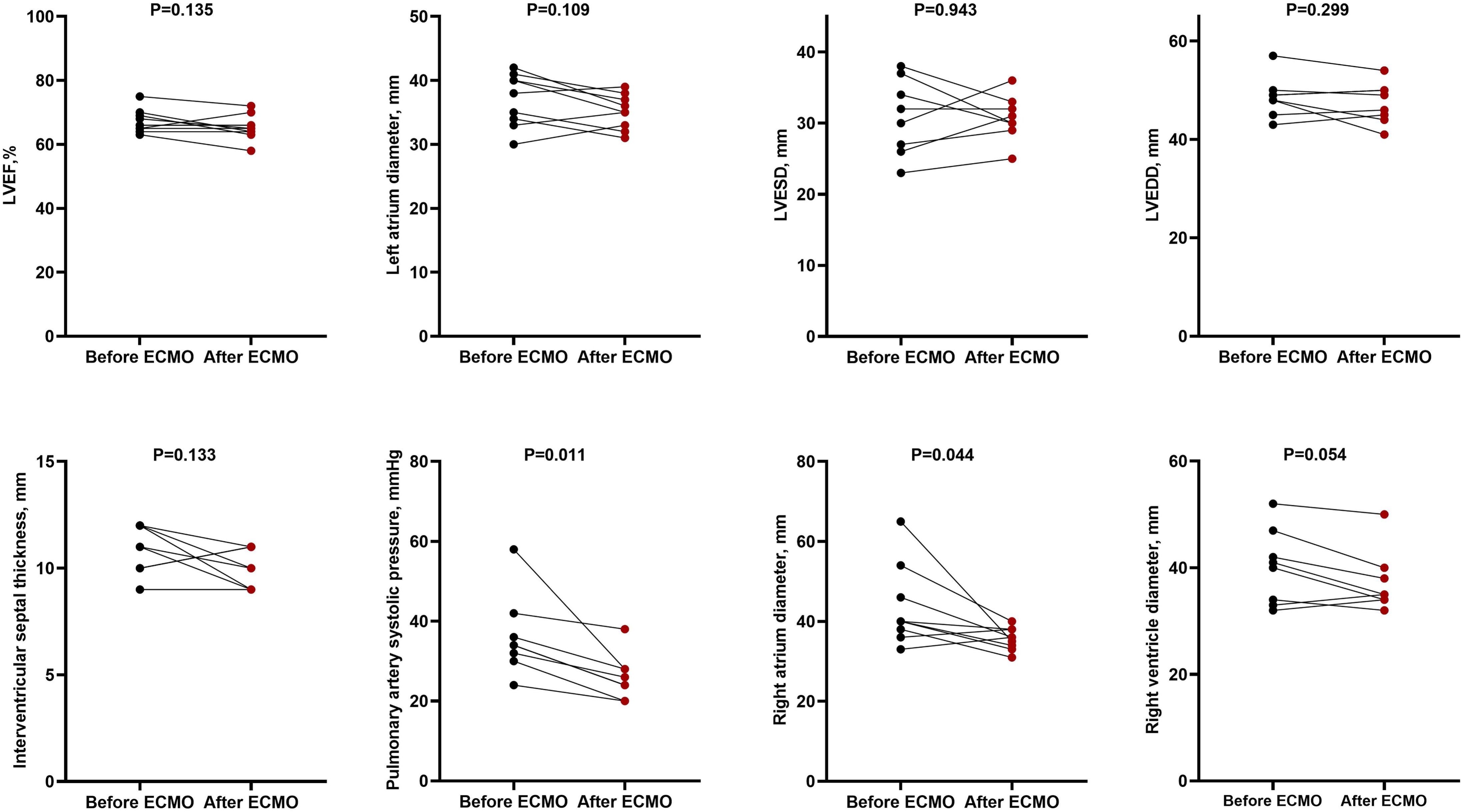
Changes in cardiac structural and functional parameters among nine COVID-19 patients treated with extracorporeal membrane oxygenation (ECMO). LVEF, left ventricular eject fraction; LVESD, left ventricular end-systolic diameter; LVEDD, left ventricular end-diastolic diameter.

### Predictors of Mortality

In univariate Cox proportional hazards regression analyses, previous malignancy, neutrophilic granulocyte percentage, lymphocyte percentage, platelet count, albumin level, hs-cTnI level, IL-6 level, IVS thickness, RA enlargement, RV enlargement, elevated PASP and decreased TAPSE were significantly associated with mortality (Table 5). Multivariable analyses identified that hs-cTnI (HR: 1.138, 95% CI: 1.029 to 1.258, *P*=0.012) and IL-6 (HR: 1.095, 95% CI: 1.002 to 1.196, *P*=0.001) were independently associated with mortality in ICU patients with COVID-19.

**Table 5.**
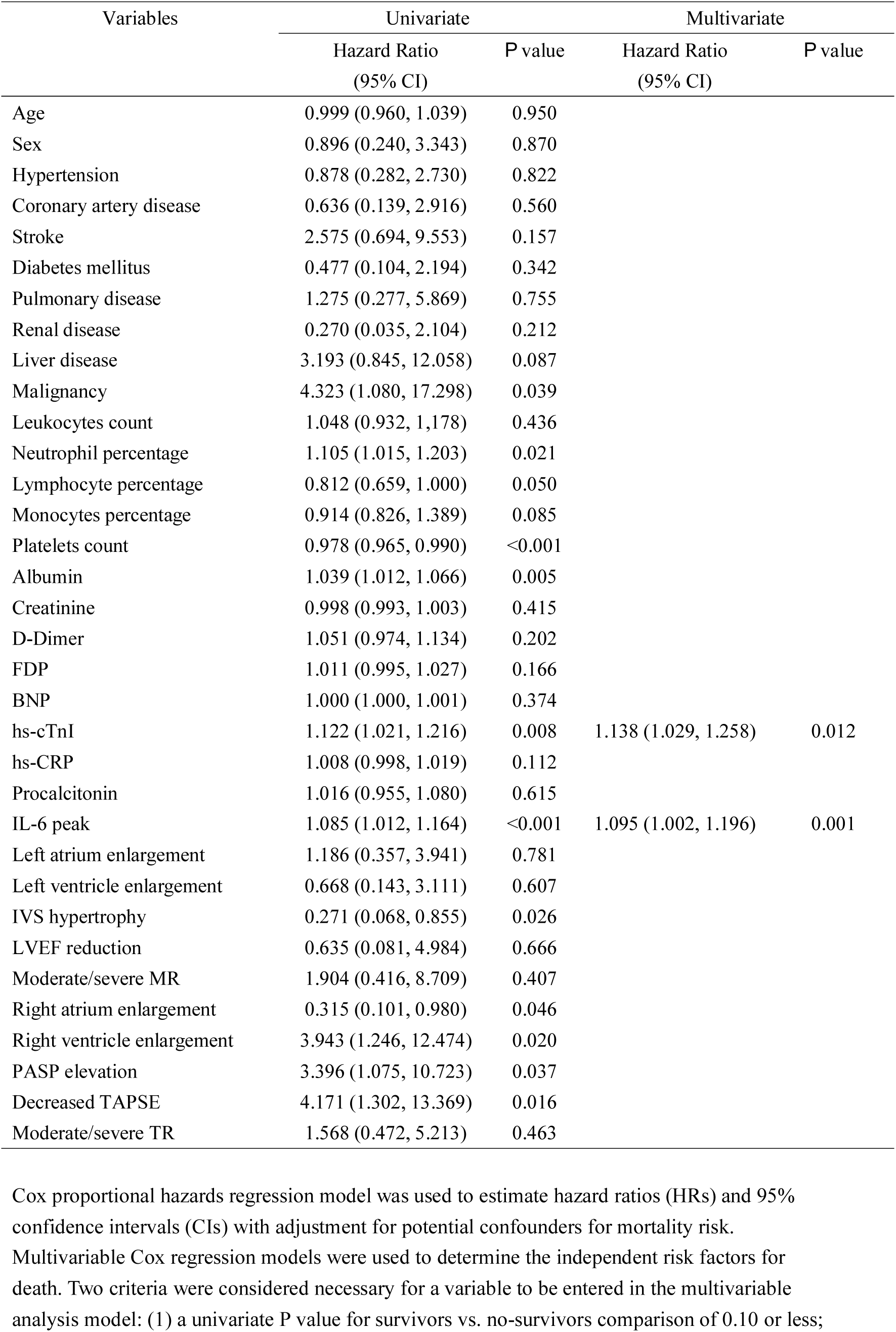
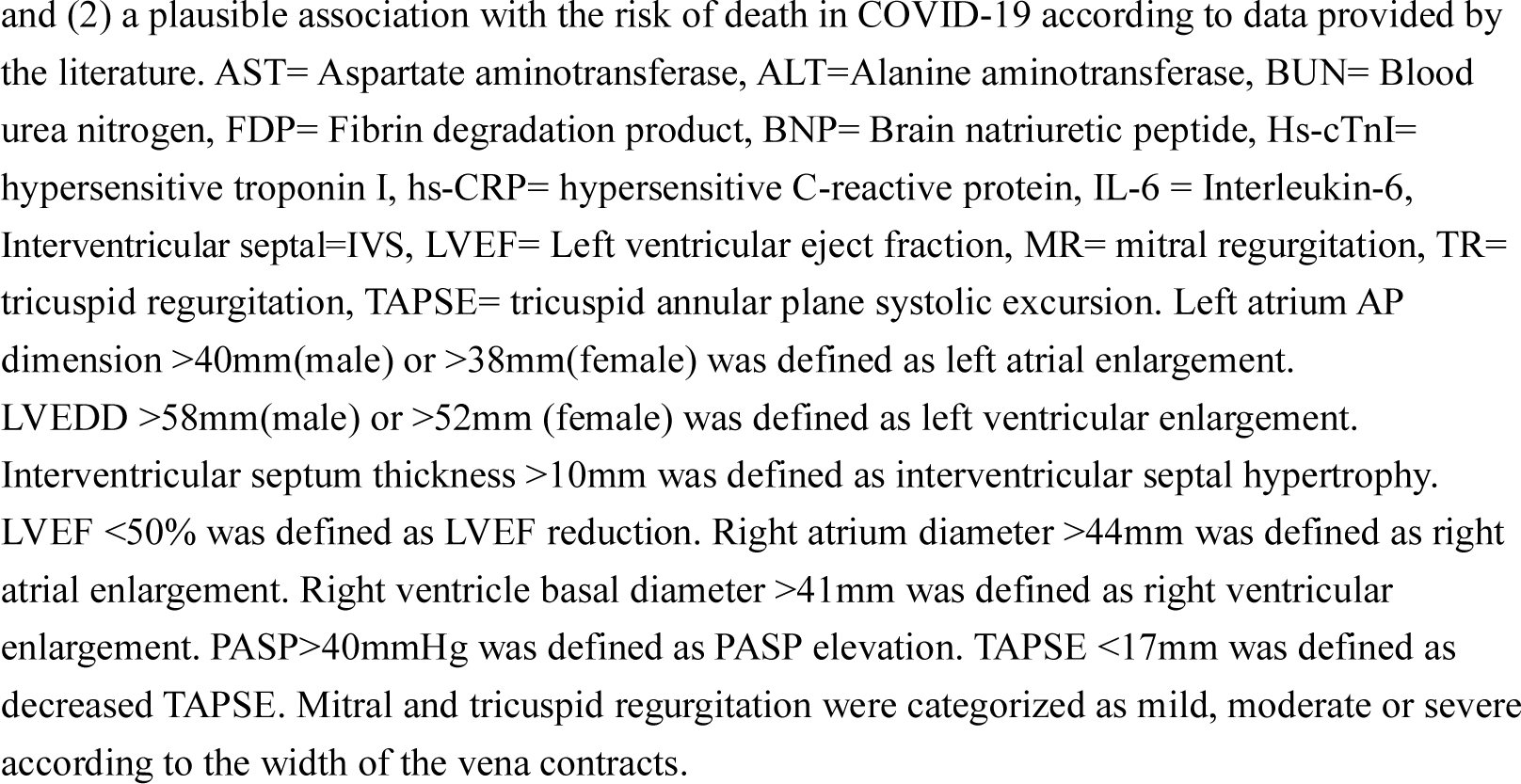
Cox Regression Analysis on the risk predictors of mortality in patients with COVID-19.

| Variables | Univariate |  | Multivariate |  |
| --- | --- | --- | --- | --- |
|  | Hazard Ratio<br>(95% CI) | *P* value | Hazard Ratio<br>(95% CI) | *P* value |
| Age | 0.999 (0.960, 1.039) | 0.950 |  |  |
| Sex | 0.896 (0.240, 3.343) | 0.870 |  |  |
| Hypertension | 0.878 (0.282, 2.730) | 0.822 |  |  |
| Coronary artery disease | 0.636 (0.139, 2.916) | 0.560 |  |  |
| Stroke | 2.575 (0.694, 9.553) | 0.157 |  |  |
| Diabetes mellitus | 0.477 (0.104, 2.194) | 0.342 |  |  |
| Pulmonary disease | 1.275 (0.277, 5.869) | 0.755 |  |  |
| Renal disease | 0.270 (0.035, 2.104) | 0.212 |  |  |
| Liver disease | 3.193 (0.845, 12.058) | 0.087 |  |  |
| Malignancy | 4.323 (1.080, 17.298) | 0.039 |  |  |
| Leukocytes count | 1.048 (0.932, 1.178) | 0.436 |  |  |
| Neutrophil percentage | 1.105 (1.015, 1.203) | 0.021 |  |  |
| Lymphocyte percentage | 0.812 (0.659, 1.000) | 0.050 |  |  |
| Monocytes percentage | 0.914 (0.826, 1.389) | 0.085 |  |  |
| Platelets count | 0.978 (0.965, 0.990) | <0.001 |  |  |
| Albumin | 1.039 (1.012, 1.066) | 0.005 |  |  |
| Creatinine | 0.998 (0.993, 1.003) | 0.415 |  |  |
| D-Dimer | 1.051 (0.974, 1.134) | 0.202 |  |  |
| FDP | 1.011 (0.995, 1.027) | 0.166 |  |  |
| BNP | 1.000 (1.000, 1.001) | 0.374 |  |  |
| hs-cTnI | 1.122 (1.021, 1.216) | 0.008 | 1.138 (1.029, 1.258) | 0.012 |
| hs-CRP | 1.008 (0.998, 1.019) | 0.112 |  |  |
| Procalcitonin | 1.016 (0.955, 1.080) | 0.615 |  |  |
| IL-6 peak | 1.085 (1.012, 1.164) | <0.001 | 1.095 (1.002, 1.196) | 0.001 |
| Left atrium enlargement | 1.186 (0.357, 3.941) | 0.781 |  |  |
| Left ventricle enlargement | 0.668 (0.143, 3.111) | 0.607 |  |  |
| IVS hypertrophy | 0.271 (0.068, 0.855) | 0.026 |  |  |
| LVEF reduction | 0.635 (0.081, 4.984) | 0.666 |  |  |
| Moderate/severe MR | 1.904 (0.416, 8.709) | 0.407 |  |  |
| Right atrium enlargement | 0.315 (0.101, 0.980) | 0.046 |  |  |
| Right ventricle enlargement | 3.943 (1.246, 12.474) | 0.020 |  |  |
| PASP elevation | 3.396 (1.075, 10.723) | 0.037 |  |  |
| Decreased TAPSE | 4.171 (1.302, 13.369) | 0.016 |  |  |
| Moderate/severe TR | 1.568 (0.472, 5.213) | 0.463 |  |  |

Cox proportional hazards regression model was used to estimate hazard ratios (HRs) and 95% confidence intervals (CIs) with adjustment for potential confounders for mortality risk.
Multivariable Cox regression models were used to determine the independent risk factors for death. Two criteria were considered necessary for a variable to be entered in the multivariable analysis model: (1) a univariate *P* value for survivors vs. no-survivors comparison of 0.10 or less;
and (2) a plausible association with the risk of death in COVID-19 according to data provided by the literature. AST= Aspartate aminotransferase, ALT=Alanine aminotransferase, BUN= Blood urea nitrogen, FDP= Fibrin degradation product, BNP= Brain natriuretic peptide, Hs-cTnI= hypersensitive troponin I, hs-CRP= hypersensitive C-reactive protein, IL-6 = Interleukin-6, Interventricular septal=IVS, LVEF= Left ventricular ejection fraction, MR= mitral regurgitation, TR= tricuspid regurgitation, TAPSE= tricuspid annular plane systolic excursion. Left atrium AP dimension >40mm(male) or >38mm(female) was defined as left atrial enlargement. LVEDD >58mm(male) or >52mm (female) was defined as left ventricular enlargement. Interventricular septum thickness >10mm was defined as interventricular septal hypertrophy. LVEF <50% was defined as LVEF reduction. Right atrium diameter >44mm was defined as right atrial enlargement. Right ventricle basal diameter >41mm was defined as right ventricular enlargement. PASP>40mmHg was defined as PASP elevation. TAPSE <17mm was defined as decreased TAPSE. Mitral and tricuspid regurgitation were categorized as mild, moderate or severe according to the width of the vena contracts.

## Discussion

The present echocardiographic study in a cohort of ICU patients with COVID-19 provided new insights into structural and functional features of cardiac involvement. The main findings were as follows: 1) Echocardiographic abnormalities were prevalent in COVID-19 patients with severe or critical condition; 2) the typical echocardiographic manifestation in COVID-19 were right-heart dysfunction with preserved LVEF; 3) right-heart abnormalities, but not left-heart abnormalities were associated with increased mortality; and 4) multivariable analyses identified hs-cTnI and IL-6 levels as independent predictors of increased mortality.

Although emerging as an acute infectious pulmonary disease, COVID-19 can cause extrapulmonary manifestations and complications^17^. Early reports included patients who died of significant heart injury ^6,11^ The fact that the COVID-19 virus invades cells through the angiotensin-converting enzyme-2 receptor, which is also abundant in the heart, raised the question of whether the virus directly targets the myocardium ^18^ In our study, increased hs-cTnI levels were observed in 62.7% of severely or critically ill ICU-admitted patients with COVID-19. This finding is consistent with previous reports of increased levels of cardiac injury biomarkers in COVID-19 ^19^ Huang et al. reported that among 41 patients with COVID-19, 31% of patients in the ICU and 4% of patients in non-ICU wards had elevated hs-cTnI level ^2^ Shi et al. found that in 416 confirmed COVID-19 patients, 19.7% had elevated hs-cTnI level, and patients with elevated hs-cTnI level had a significantly higher mortality ^4^. The exact mechanism underlying cardiac injury caused by COVID-19 remains unclear although speculation includes a mechanistic link to hyperinflammation/cytokine storm ^20,21^. In the present study, severe systemic inflammatory stress was observed among ICU patients with COVID-19, as evidenced by significantly elevated hs-CRP and IL-6 levels. Moreover, we identified both hs-cTnI and IL-6 as predictors of mortality.

### Left Heart Abnormalities

Lessons from previous coronavirus and influenza epidemics suggested that the virus would either induce new cardiac pathologies and/or exacerbate underlying cardiovascular diseases ^22^. It was postulated that the systemic inflammatory storm in COVID-19 might lead to acute myocarditis^6^, stress cardiomyopathy ^23^, or acute coronary syndromes ^7^, recently described as the “acute COVID-19 cardiovascular syndrome” ^24^ The latter should significantly impair LV systolic function, which might be expected to be a key characteristic of cardiac involvement of patients with COVID-19. LVEF has been confirmed as a key determinant of prognosis in cardiac patients ^25,26^. Therefore, it was surprising and unexpected to find that in our cohort of severely and critically ill COVID-19 patients, the majority had preserved LVEF; only 15.7% had reduced LVEF that was often associated with pre-existing heart disease. Moreover, LVEF was comparable in critically ill vs. severely ill COVID-19 patients and in COVID-19 survivors vs. non-survivors; serial echocardiography did not reveal LVEF deterioration in non-survivors from ICU admission to death; and left-heart abnormalities were not associated with increased mortality in COVID-19 patients. Thus, our findings did not support the notion that LV systolic dysfunction was the principal form of cardiac involvement associated with COVID-19.

In contrast, we observed abnormal echocardiographic manifestations associated with impaired LV diastolic function in COVID-19 patients, as evidenced by E/A ratio and E/e’ ratio abnormality and IVS hypertrophy. The impaired LV diastolic function could not be attributed to the underlying cardiovascular or other comorbidities as the comorbidities were comparable in critically ill vs. severely ill patients as well as in non-survivors vs. survivors. Moreover, IVS thickness paralleled disease severity and even increased significantly in non-survivors from ICU admission to death. Similarly, a previous study found that subclinical diastolic left ventricular impairment was common in acute severe acute respiratory syndrome (SARS) infection^27^.

Our findings of impaired LV diastolic function with preserved LV systolic function were in accordance with the findings of several recent autopsy studies in COVID-19 ^8,28^. On one hand, no apparent myocardial necrosis/apoptosis was found in myocardium in COVID-19 at autopsy ^8,28^. Consistently, although the prevalence was high, the elevation of hs-cTnI level was mostly mild (median 0.07 [IQR 0.02–0.23]) in our cohort. This may explain the preserved LVEF among patients studied. On the other hand, the diastolic LV dysfunction in the current study was similar to that in Clancy’s report in which as many as 60% of patients with severe sepsis had echocardiographic manifestations of diastolic dysfunction ^29^ Furthermore, several studies have reported that diastolic rather than systolic LV dysfunction was an independent predictor of mortality in patients with severe sepsis ^30,31^. Xu et al. reported a patient who died from severe COVID-19 who had mononuclear inflammatory infiltration in the myocardial interstitium, but no substantial damage to the myocytes. Tian et al. reported four patients who died of COVID-19 who had various degrees of cardiac focal edema and interstitial fibrosis ^28^. Inciardi et al. described cardiac magnetic resonance imaging showing marked ventricular interstitial edema and increased wall thickness in a 53-year-old woman with COVID-19^6^. Taking together, the increased IVS thickness and deteriorating LV compliance observed in our study might be reflective of interstitial inflammatory edema, as has been found in other sepsis studies ^29^.

### Right Heart Abnormalities

About one third of the study patients had right-heart abnormalities as evidenced by elevated PASP, larger RA and RV diameters, and decreased TAPSE. Among them, only 13.6 % had a history of chronic lung disease. Moreover, right-heart dysfunction, but not left-heart dysfunction, was associated with increased mortality in our cohort. The mechanism of right-heart dysfunction in COVID-19 is unclear but may be related to vascular thickening in the lung ^32^, hypoxemia and pro-inflammatory cytokines that would provoke pulmonary vasoconstriction ^2^, or the possibility that pulmonary embolism as suggested by the extremely high concentration of D-dimer (observed in our cohort and other populations) and its close relationship with mortality ^33^. A postmortem study in eight patients who died from SARS showed that four patients had pulmonary thromboembolism ^34^. Rapid development and progression of right-heart dysfunction has been reported in patients with acute respiratory distress syndrome or acute pulmonary embolism ^35^. The adverse impact of right-heart impairment on prognosis also was reported in ARDS patients ^36,37^

### Impact of ECMO

Reports of ECMO utility in COVID-19 patients have been scarce, and its clinical effectiveness and outcomes remained unknown. We monitored the dynamic changes of cardiac structural and functional parameters before and after ECMO implantation. ECMO implantation tended to alleviate elevated PASP and other right-heart dysfunction in critically ill patients. Theoretically, the application of ECMO might reduce pulmonary hypertension by improving oxygenation, correcting hypoxemia and alleviating pulmonary vasoconstriction. Moreover, RV volume may be unloaded with a venoarterial ECMO. However, in non-survivors treated with ECMO, the echocardiographic improvement was temporary; and the right-heart dysfunction deteriorated again before death (Supplementary Figure 1), similar to the post ECMO scenario in children with pulmonary hypertension from pertussis ^38^. Thus, it seems that the key value of ECMO implantation is as a temporary life support to gain time for therapy, and the treatment of the primacy pulmonary disease is vital for the final prognosis.

## LIMITATIONS

First, although data in our study were prospectively collected and analyzed in a blinded fashion, the sample size was small. Second, new echocardiographic technologies (i.e., speckle tracking technology for global and regional function and deformation) and cardiac magnetic resonance imaging (MRI) were not used. The fact that the current study included ICU patients on mechanical respiratory and/or circulatory support and strong infectious inherence limited the application of these new imaging technologies. Third, data were obtained from severely or critically ill ICU patients and may not be representative of the general population.

## CONCLUSIONS

Right-heart dysfunction with preserved LVEF is the typical characteristic of cardiac involvement in ICU patients with COVID-19. Moreover, right-heart, but not left-heart dysfunction was associated with disease severity and increased mortality. Our findings will facilitate unraveling the underlying mechanisms of myocardial injury and optimize treatment strategy for patients with COVID-19.

## Data Availability

All epidemiological, clinical, laboratory, and outcome parameters were prospectively collected with standardized data collection forms from an electronic medical records system.

## ACKNOWLEDGMENTS

We are grateful to Dr. Gary Mintz for his critical and constructive discussion, and manuscript refinement.

